# United Network for Organ Sharing Policy Change Impact on Competing Risks in Heart Transplantation: Landmark Analysis of Waitlist Outcomes over Time

**DOI:** 10.64898/2026.09.29.26362912

**Authors:** Aaron M. Wolfson, Matthew S. Chen, Joel Ferrall, Jonathan Kermanshahchi, Michael W. Fong, Jonathan Nattiv, Maxine Stachel, Eric S. Kawaguchi, Ajay S. Vaidya

**Author notes:** **Address for Correspondence:** Aaron M. Wolfson, MD, MS, 1510 San Pablo Street, Suite 322, Los Angeles, CA 90033, Academic, Twitter: @wolfsonaaron. co-first authors (Aaron M. Wolfson, Matthew S. Chen).

## Abstract

**Background:** Studies of the 2018 United Network for Organ Sharing (UNOS) policy revision suggest improved early transplantation access for the sickest patients. It remains unknown whether these benefits vary as a function of time on the waitlist. Thus, we evaluated potential time-dependent effects of the 2018 heart allocation policy revision on waitlist outcomes.

**Methods:** Using UNOS data, we analyzed adult heart transplant candidates listed five years before (pre-policy) and after (post-policy) policy change, categorized as high-acuity (Status 1-3 or 1A) or low-acuity (Status 4-6 or 1B/old 2). Landmark analysis at weeks 0, 1, 2, 4, 8, 16, 24 using a competing risks framework was performed to assess transplantation, death/deterioration, and myocardial recovery. Sub-distribution and cause-specific hazard ratios were calculated. Absolute number and proportion of far-away transplants (donor distance ≥500 nautical miles) were calculated at each timepoint.

**Results:** High-acuity patients experienced significantly higher risk of transplantation within 2 weeks post-policy change but equivocal risk by week 16. Low-acuity patients exhibited increased rate/risk of myocardial recovery and equal or improved death/deterioration risk at all timepoints; early transplantation cumulative incidence was increased. At all timepoints, both groups experienced significantly increased far-away donor use, highest immediately after listing and progressively declining over time.

**Conclusions:** The 2018 heart allocation policy change resulted in an initial appearance of time-dependent benefits among high-acuity, post-policy patients, but this was likely due to rapid transplantation of Status 1 and 2 patients from broader geographic sharing rather than a true temporal phenomenon. Post-policy, low-acuity patients experience benefits in all waitlist outcomes.

## 1.0 Introduction

Heart transplantation remains the definitive therapy for selected patients with end-stage heart failure (HF), but donor allocation continues to evolve [1]. The 2018 revision to the United Network of Organ Sharing (UNOS) heart allocation policy system introduced a six-tiered scheme and broader geographic sharing to prioritize donor organ availability to the highest-acuity candidates [2]. Specifically, broader geographic sharing expanded access of donor hearts to 500 nautical miles (NM) from the recipient hospital for Status 1 and 2 patients [3]. Early studies show that broader sharing was associated with increased transplantation over greater distances, improved waitlist outcomes in the sickest candidates, and more balanced donor distribution [4–7]. Other studies reported reduced waitlist mortality [8–10] and higher transplantation rates in select patient subgroups [2, 11]. However, nearly all of these analyses failed to incorporate time on the waitlist or update patients’ evolving listing statuses, implicitly assuming that the effects of the revised allocation system remain constant throughout a patient’s waitlist course.

Several observations suggest that this assumption may be incorrect. Prior competing risk studies show early separation of cumulative incidence curves for transplantation and waitlist mortality between eras followed by attenuation over time [12, 13]. These patterns could reflect true temporal changes in risk, but they may also arise from shifts in waitlist composition— particularly early removal of high-acuity candidates through transplantation and the accumulation or upgrade of lower-acuity candidates over time. Traditional analytic methods do not account for this dynamic risk-set evolution, limiting the ability to determine whether observed differences represent a true time-dependent policy effect or the consequences of a changing waitlist population. Understanding whether the revised allocation system exerts different effects at different intervals after listing is important for contemporary transplant practice (i.e. when to consider durable left ventricular assist device (LVAD) implantation) and future policy iterations. This is especially important as we move toward a continuous distribution model for heart allocation. Temporal effects on risk may prove to be a factor when delineating allocation priority.

To address this knowledge gap, we evaluated the time-dependent effects of the 2018 UNOS allocation policy on transplantation, waitlist death/deterioration, and myocardial recovery using a landmark competing-risks framework stratified by clinical acuity.

## 2.0 Material and Methods

### Database and Patient Cohort

Following University of Southern California Institutional Review Board exemption, we conducted a retrospective analysis using the UNOS registry. Adult candidates (≥18 years old) listed for first-time heart, heart–kidney, or heart–liver transplantation were included; prior heart transplantation or other multi-organ transplantation types were excluded.

Two cohorts were categorized based on the October 18, 2018 policy revision implementation date. The pre-policy cohort included patients listed between October 18, 2012–August 30, 2017, and the post-policy cohort included patients listed between October 18, 2018–August 30, 2023. Candidates listed between September 1, 2017–October 17, 2018 were excluded as a washout period. Follow-up was administratively censored on September 30, 2018 (pre-policy) and on September 30, 2024 (post-policy), providing similar follow-up waitlist durations for both groups.

To harmonize acuity definitions across policy eras, patients were classified as high-acuity or low-acuity based on their listing status at each landmark time. In the pre-policy cohort, high-acuity candidates were those listed as Status 1A, and low-acuity candidates were those listed as Status 1B or Status 2 under the previous allocation system. In the post-policy cohort, high-acuity patients were defined as those listed under Status 1, 2, or 3, while low-acuity patients were those listed under Status 4, 5, or 6. Because veno-arterial extracorporeal membrane oxygenation (VA-ECMO), intra-aortic balloon pump (IABP), VAD, or mechanical ventilation is incompatible with low-acuity listing, 656 “low-acuity” candidates who received these forms of advanced support were excluded.

### Exposure and Outcomes

The primary exposure was listing in the pre-policy versus post-policy era. The outcomes of interest were delineated as competing risks, namely heart transplantation, death or clinical deterioration leading to delisting, and myocardial recovery leading to delisting. Individuals delisted for other causes were considered right censored. Outcomes were evaluated at prespecified landmark times of 0, 1, 2, 4, 8, 16, and 24 weeks following initial listing date.

### Statistical Analysis

Baseline characteristics were summarized using descriptive statistics. Continuous variables were reported as medians with interquartile ranges, and categorical variables were summarized as frequencies with percentages. To compare groups, the Kruskal–Wallis test for continuous variables and the chi-square test for categorical variables were used.

Competing risks analyses were used to estimate subdistribution hazard ratios (sdHRs) and cause-specific hazard ratios (csHRs) for each outcome. The cumulative incidence of each event was compared using the Fine–Gray proportional subdistribution hazards model [14], whereas cause-specific Cox proportional hazards models evaluated the cause-specific hazard of each outcome, both adjusting for age, sex, race or ethnicity, blood type, and body mass index.

### Landmark Analysis

To investigate potential time-varying effects of listing era and to incorporate updated listing status during the waitlist course, we performed a landmark analysis [15]. This evaluates the association between listing era and waitlist outcomes at prespecified intervals after listing. Patients who already experienced an outcome or were lost to follow-up before a given landmark time were excluded from that risk set. Landmark times were selected at 0, 1, 2, 4, 8, 16, and 24 weeks to provide adequate temporal resolution to detect early and late policy effects. At each landmark, listing status was reassigned using the most recent listing data available prior to that timepoint, followed by competing risks analysis to estimate sdHR and csHR for each outcome. A landmark analysis allows for individual patients to be reassigned into a status as mechanical or pharmacological interventions are used or discontinued in the waitlist course.

In a separate analysis assessing changes in donor geography, we evaluated the number and proportion of transplant procedures at each landmark time involving donor hearts procured ≥500 nautical miles (NM) from the recipient. These cases were classified as “far-away” donor utilization. A two-proportion Z-test was used to compare proportions between groups.

All statistical analyses were conducted using R (version 4.2.0, R Foundation for Statistical Computing). A two-sided p-value < 0.05 was considered statistically significant.

## 3.0 Results

### Baseline Clinical Characteristics

Among 35,520 included candidates, 12,320 were high-acuity (4,121 pre-policy change, 8,199 post-policy change) and 23,200 were low-acuity (12,236 pre-policy change, 10,964 post-policy change). Baseline patient characteristic data are available in Tables 1 and 2.

**Table 1.** Baseline clinical characteristics of heart transplant candidates stratified by acuity status.

| Low-acuity patients |  |  |  | High-acuity patients |  |  |
| --- | --- | --- | --- | --- | --- | --- |
| Variable | Pre-policy<br>change,<br>N = 12,236 | Post-policy<br>change,<br>N = 10,964 | p-value | Pre-policy<br>change,<br>N = 4,121 | Post-policy<br>change,<br>N = 8,199 | p-value |
| Age | 56 (47-63) | 57 (47-63) | 0.031 | 55 (45-62) | 56 (44-63) | 0.388 |
| Sex, Male | 9105 (74.4%) | 7981 (72.8%) | 0.005 | 3099 (75.2%) | 6214<br>(75.8%) | 0.486 |
| BMI | 27.80 (24.4-31.5) | 28.46 (25.0-32.3) | <0.001 | 27.12 (23.6-30.9) | 27.12 (23.6-31.2) | 0.202 |
| Diabetes mellitus | 3698 (30.2%) | 3430 (31.3%) | 0.079 | 1273 (30.9%) | 2473<br>(30.2%) | 0.409 |
| Ischemic<br>cardiomyopathy<br>* | 4258 (34.8%) | 3362 (30.7%) | <0.001 | 1416 (34.4%) | 2175<br>(26.5%) | <0.001 |
| Prior cardiac<br>surgery | 5069 (41.4%) | 4584 (41.8%) | 0.551 | 1777 (43.2%) | 2578<br>(31.5%) | <0.001 |
| ICD | 9850 (80.5%) | 8083 (73.8%) | <0.001 | 2981 (72.4%) | 4984<br>(60.8%) | <0.001 |
| Race/Ethnicity |  |  | <0.001 |  |  | <0.001 |
| Asian | 373 (3.0%) | 319 (2.9%) |  | 162 (3.9%) | 382 (4.7%) |  |
| Black | 2855 (23.3%) | 2831 (25.8%) |  | 1051 (25.5%) | 2254<br>(27.5%) |  |
| Hispanic | 1011 (8.3%) | 1053 (9.6%) |  | 372 (9.0%) | 956 (11.7%) |  |
| Other | 0 (0.0%) | 0 (0.0%) |  | 0 (0.0%) | 0 (0.0%) |  |
| White | 7875 (64.4%) | 6628 (60.5%) |  | 2472 (60.0%) | 4496<br>(54.8%) |  |
| AI/AN | 37 (0.3%) | 49 (0.4%) |  | 19 (0.5%) | 35 (0.4%) |  |
| NH/PI | 43 (0.4%) | 30 (0.3%) |  | 25 (0.6%) | 35 (0.4%) |  |
| Multiracial | 42 (0.3%) | 54 (0.5%) |  | 20 (0.5%) | 41 (0.5%) |  |
| Missing | 0 | 0 |  | 0 | 0 |  |
| **Dialysis at registration** | 270 (2.2%) | 433 (4.0%) | <0.001 | 149 (3.6%) | 393 (4.8%) | 0.003 |
| **Mechanical ventilation at registration** | 0 | 0 |  | 174 (4.2%) | 305 (3.7%) | 0.19 |
| **IV inotropes** | 3401 (27.8%) | 2271 (20.7%) | <0.001 | 1676 (40.7%) | 3756 (45.8%) | <0.001 |
| **IABP** | 0 | 0 |  | 632 (15.3%) | 2473 (30.2%) | <0.001 |
| **VA-ECMO** | 0 | 0 |  | 201 (4.9%) | 679 (8.3%) | <0.001 |
| **Number of organs waitlisted for** |  |  | <0.001 |  |  | <0.001 |
| **One** | 11353 (92.8%) | 9556 (87.2%) |  | 3779 (91.7%) | 7129 (86.9%) |  |
| **Two** | 883 (7.2%) | 1408 (12.8%) |  | 342 (8.3%) | 1070 (13.1%) |  |
| **VAD Type** |  |  | 0.585 |  |  | <0.001 |
| **None** | 8836 (72.2%) | 7951 (72.6%) |  | 2409 (58.5%) | 5644 (68.9%) |  |
| **LVAD** | 3398 (27.8%) | 3008 (27.4%) |  | 1466 (35.6%) | 2288 (27.9%) |  |
| **RVAD** | 0 (0.0%) | 0 (0.0%) |  | 9 (0.2%) | 35 (0.4%) |  |
| **TAH** | 0 (0.0%) | 0 (0.0%) |  | 63 (1.5%) | 38 (0.5%) |  |
| **BiVAD** | 0 (0.0%) | 0 (0.0%) |  | 171 (4.2%) | 192 (2.3%) |  |
| **Missing** | 2 | 5 |  | 3 | 2 |  |
| **Initial listing status** |  |  | <0.001 |  |  | <0.001 |
| **1** | 0 (0.0%) | 0 (0.0%) |  | 0 (0.0%) | 957 (11.7%) |  |
| **2** | 0 (0.0%) | 0 (0.0%) |  | 0 (0.0%) | 5119 (62.4%) |  |
| **3** | 0 (0.0%) | 0 (0.0%) |  | 0 (0.0%) | 2123 (25.9%) |  |
| **4** | 0 (0.0%) | 6702 (61.1%) |  | 0 (0.0%) | 0 (0.0%) |  |
| **5** | 0 (0.0%) | 507 (4.6%) |  | 0 (0.0%) | 0 (0.0%) |  |
| **6** | 0 (0.0%) | 3755 (34.2%) |  | 0 (0.0%) | 0 (0.0%) |  |
| **1A** | 0 (0.0%) | 0 (0.0%) |  | 4121 (100.0%) | 0 (0.0%) |  |
| **1B** | 7661 (62.6%) | 0 (0.0%) |  | 0 (0.0%) | 0 (0.0%) |  |
| **2A** | 4575 (37.4%) | 0 (0.0%) |  | 0 (0.0%) | 0 (0.0%) |  |

**Table 2.** Baseline hemodynamics stratified by patient acuity.

| Hemodynamic variable | Low-acuity patients |  |  | High-acuity patients |  |  |
| --- | --- | --- | --- | --- | --- | --- |
|  | Pre-policy change, N = 12,236 | Post-policy change, N = 10,964 | p-value | Pre-policy change, N = 4,121 | Post-policy change, N = 8,199 | p-value |
| **Pulmonary artery systolic pressure (mm Hg)** | 41.00 (31.0-52.0) | 36.00 (28.0-47.0) | <0.001 | 43.00 (33.0-53.0) | 44.00 (34.0-54.0) | 0.028 |
| **Pulmonary artery diastolic pressure (mm Hg)** | 20.00 (14.0-25.0) | 17.00 (12.0-23.0) | <0.001 | 22.00 (16.0-28.0) | 22.00 (16.0-28.0) | 0.019 |
| **Mean pulmonary artery pressure (mm Hg)** | 28.00 (21.0-35.0) | 24.00 (18.0-32.0) | <0.001 | 30.00 (22.2-37.0) | 30.00 (23.0-38.0) | 0.022 |
| **Pulmonary capillary wedge pressure (mm Hg)** | 18.00 (12.0-25.0) | 15.00 (10.0-22.0) | <0.001 | 20.00 (14.0-27.0) | 21.00 (15.0-28.0) | <0.001 |
| **Cardiac Index (L/min/m<sup>2</sup>)** | 2.12 (1.8-2.5) | 2.12 (1.8-2.5) | 0.271 | 2.10 (1.7-2.6) | 2.01 (1.6-2.5) | <0.001 |

In the high-acuity cohort, ischemic cardiomyopathy prevalence (26.5% vs. 34.4%, p < 0.001), implantable cardioverter-defibrillator (ICD) use (60.8% vs. 72.4%, p < 0.001), and VAD use (no VAD: 58.5% vs. 68.9%, p < 0.001) were lower post-policy. Intravenous inotrope (45.8% vs. 40.7%, p < 0.001), IABP (30.2% vs. 15.3%, p < 0.001), and VA-ECMO (8.3% vs. 4.9%, p < 0.001) usage increased post-policy. More high-acuity patients were listed for two organs post-policy (13.1% vs. 8.3%, p < 0.001).

Among low-acuity patients, there was a reduction in male candidates (72.8% vs. 74.4%, p = 0.005) and ischemic cardiomyopathy (30.7% vs. 34.8%, p < 0.001) post-policy. More patients post-policy required dialysis (4.0% vs. 2.2%, p < 0.001), while fewer used an ICD (73.8% vs. 80.5%, p < 0.001) or intravenous inotropes (20.7% vs. 27.8%, p < 0.001). The proportion of two-organ listings increased (12.8% vs. 7.2%, p < 0.001).

Over time, the total number and proportion of Status 1 and 2 patients declined rapidly relative to Status 3 patients: by week 16, there is a greater proportion of Status 3 patients compared to Status 1 or 2 (Figure 1A). Conversely, the proportions of Status 4, 5 and 6 patients remain stable over time (Figure 1B).

**Figure 1.**
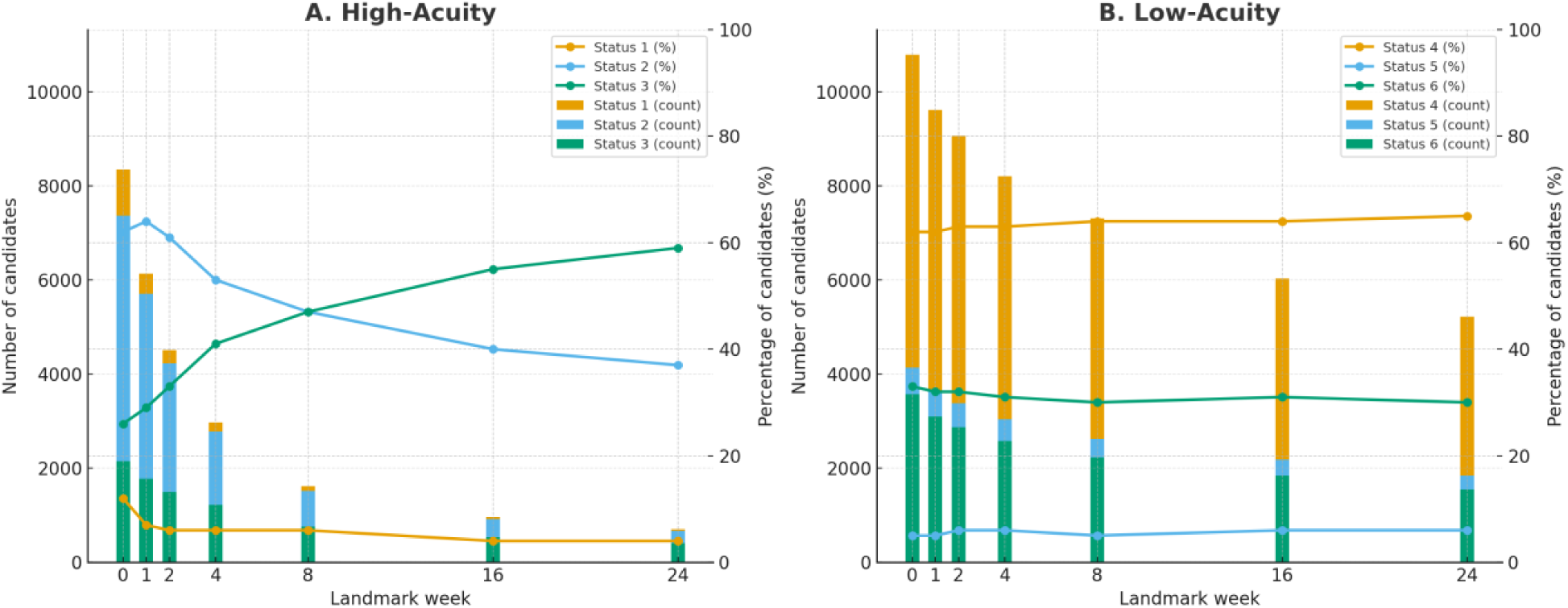
Proportions of patients based on status classification in the study population over time for high-acuity patients (left side, A) vs. low-acuity patients (right side, B).

### Waitlist Outcomes

#### Heart Transplantation

In high-acuity patients (Figure 2A, 2C; Supplemental Table 1), the sdHR for heart transplantation was initially significantly higher post-policy but decreased over time. At week 0, the sdHR was 2.13 (95% CI: 2.04, 2.22; p < 0.001), and by week 16 the sdHR dropped below 1 (0.89, 95% CI: 0.81, 0.99; p = 0.027). Similarly, the csHR started at 2.29 (95% CI: 2.19, 2.38; p < 0.001) at week 0 and declining to 0.87 (95% CI: 0.79, 0.96; p = 0.0052) by week 16.

**Figure 2.**
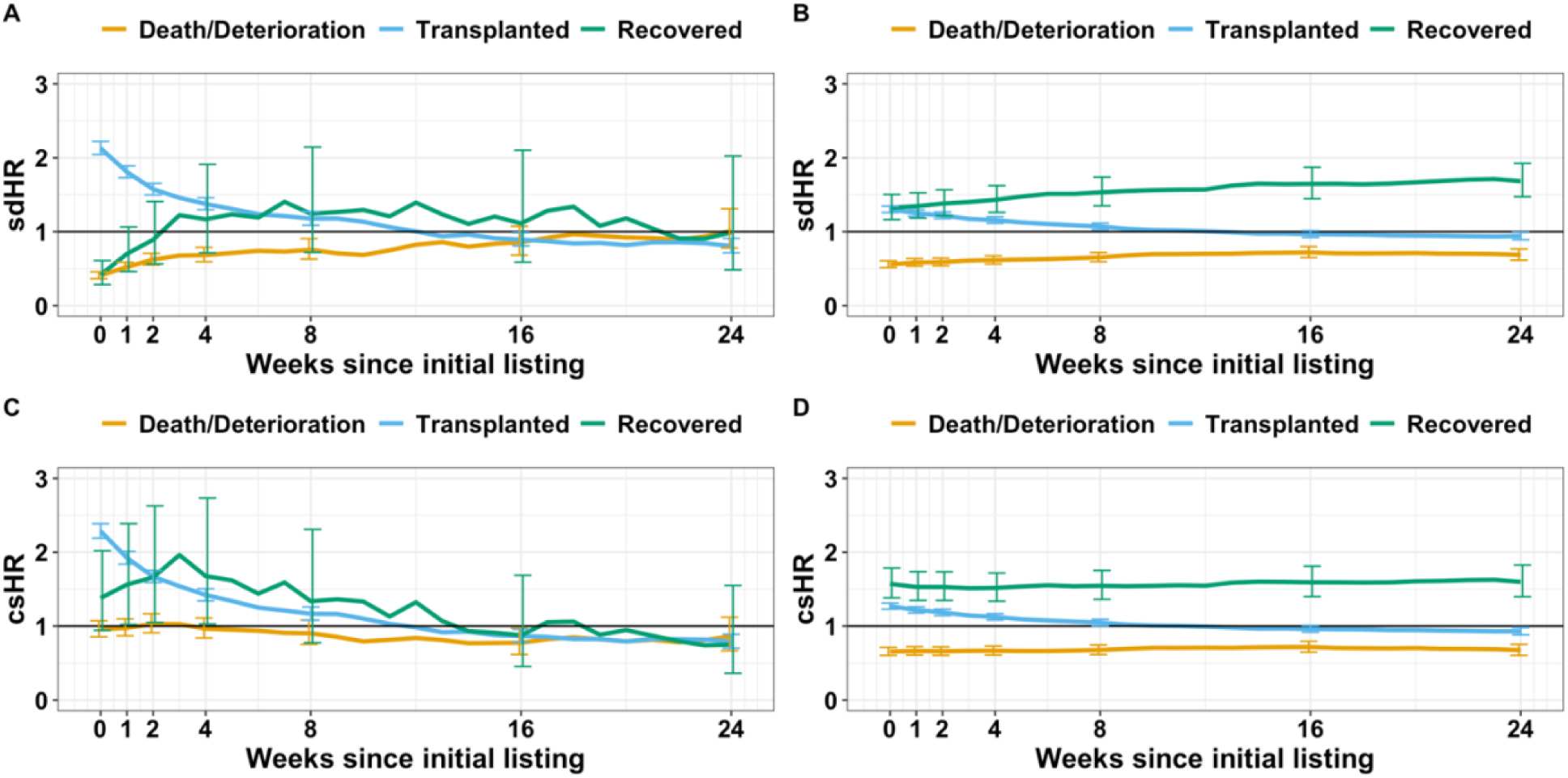
Landmark Analysis at multiple time points for high-acuity patients (left side, A and C) vs. low-acuity patients (right side, B and D).

For low-acuity patients (Figure 2B, 2D; Supplemental Table 2), the sdHR for heart transplantation was initially elevated in the post-policy group (sdHR 1.30, 95% CI: 1.26, 1.35; p < 0.001 at week 0) but gradually decreased, reaching 0.94 (95% CI: 0.89, 0.99; p = 0.022) at week 24. The csHR was also initially increased (1.27, 95% CI: 1.23, 1.31; p < 0.001) and declined to 0.93 (95% CI: 0.88, 0.98; p = 0.008) by week 24.

#### Death or Clinical Deterioration

For high-acuity patients, the risk of death or deterioration was lower post-policy at week 0 (sdHR 0.41, 95% CI: 0.37, 0.46; p < 0.001), a protective effect that persisted but diminished by week 24 (sdHR 1.01, 95% CI: 0.78, 1.31; p = 0.93. The csHR was not significantly different at most time points, though it was lower in the post-policy group at week 16 (csHR 0.78, 95% CI: 0.62, 0.98; p = 0.03).

For low-acuity patients, death or deterioration risk was significantly lower post-policy at all timepoints. The sdHR was 0.56 (95% CI: 0.51, 0.61; p < 0.001) at week 0 and 0.69 (95% CI: 0.62, 0.77; p < 0.001) by week 24. The csHR was also consistently lower post-policy with similar reductions in rate over time.

#### Myocardial Recovery

Myocardial recovery likelihood was lower post-policy at week 0 (sdHR 0.42, 95% CI: 0.29, 0.61; p < 0.001) in high-acuity patients but was not significant beyond this. The csHR for myocardial recovery was initially not significant (p = 0.097 week 0) but significant at weeks 2 (csHR 1.66, 95% CI: 1.05, 2.63; p = 0.030) and 4 (csHR 1.68, 95% CI: 1.03, 2.73; p = 0.038). The csHR was not significant at later timepoints.

For low-acuity patients, myocardial recovery likelihood was higher post-policy at all timepoints. The sdHR increased from 1.32 (95% CI: 1.17, 1.50; p < 0.001) at week 0 to 1.68 (95% CI: 1.47, 1.93; p < 0.001) at week 24. Similarly, the csHR remained significantly elevated across all timepoints.

#### Changes in Usage of Far-Away Donors over Time

Figure 3A shows that among high-acuity patients post-policy change, there was significantly increased far-away donor usage at each landmark, both in total number and overall proportion of transplanted recipients (p < 0.001 weeks 1-24). Post-policy, the number of far-away recipients decreased from 491 at week 1 to 58 by week 24; the proportion decreased from 17% at week 1 to 11% by weeks 8 and 16. Conversely, pre-policy high-acuity patients had similar counts from week 1-24, ranging between 27-47 with proportions between 4-8%.

**Figure 3.**
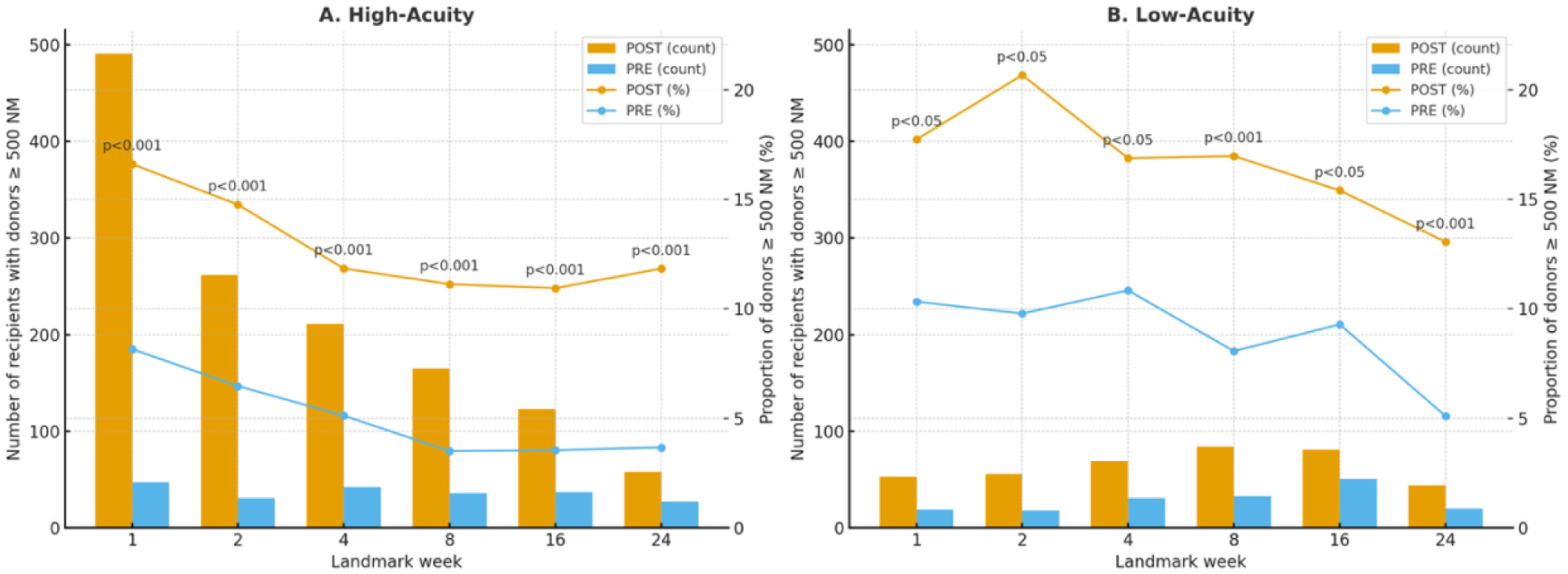
Total number of transplant recipients (bar graph, left-sided axes) and proportion of overall transplanted patients (line graph, right-sided axes) receiving hearts from donors ≥500 NM away from the recipient at different landmark time points. Separated into high-acuity (left side, A) and low-acuity (right side, B). p-values for differences in proportion between PRE (pre-policy change) and POST (post-policy change).

Similarly, Figure 3B demonstrates significantly higher usage of far-away donors among low-acuity patients at all timepoints post-policy change, both in total number and proportion (p < 0.05 weeks 1-4, 16; p < 0.001 weeks 8, 24). At weeks 1 and 2 post-policy change, low-acuity patients had proportions of far-away donor usage of 18% and 21%, respectively, that decreased to 13% by week 24. Pre-policy change, far-away donor usage rate decreased from 10% in week 1 and 2 to 5% by week 24. Further information on far-away donor usage is found in Supplemental Table 3.

## 4.0 Discussion

In our analysis using a landmark competing-risks framework, we evaluated whether the 2018 UNOS heart allocation policy change exerted time-dependent effects on waitlist outcomes. We made several key observations. First, among high-acuity patients, the post-policy era was characterized by a marked early increase in the rate and cumulative incidence of transplantation, followed by a progressive decline that became neutral or even unfavorable relative to the pre-policy era by 16-24 weeks. Second, this apparent “time-dependent” pattern coincided with rapid early removal of Status 1 and 2 candidates from the risk set and with the highest rates of long-distance donor utilization occurring shortly after listing. Third, in contrast to high-acuity patients, post-policy low-acuity patients experienced favorable changes across all waitlist outcomes (early heart transplantation, death/deterioration and myocardial recovery) throughout follow-up. Finally, compared to the pre-policy era, both high- and low-acuity patients post-policy demonstrated increased utilization of far-away donors.

The large increase in early post-policy transplantation hazard among high-acuity patients in our analysis is aligned with the intended goal of the 2018 allocation policy change: accelerating organ access for the sickest candidates through broader geographic sharing and acuity-based distribution [2–7, 16]. At listing, high-acuity patients in the post-policy era had more than twice the rate and incidence of transplantation compared with pre-policy counterparts, but this effect waned quickly as Status 1 and 2 patients were transplanted and removed from the risk set. By week 16, transplantation rates were no longer elevated and ultimately fell below those observed in the pre-policy era.

Our findings suggest that the observed decline in heart transplant hazard ratios does not necessarily represent a diminished policy effect over time but instead reflects the rapid depletion of the most urgent patients from the waitlist as donor hearts become available. The hazard estimates later in the listing course may represent a less critically ill population where urgency of donor acceptance is lower than the status-assigned patient acuity. This interpretation is reinforced by the parallel decline in far-away donor utilization over time. The magnitude and proportion of long-distance donor use was highest immediately after listing then dropped sharply. This is consistent with the optimization of broader geographic sharing during the period in which the highest-acuity patients were still present on the waitlist [4, 5, 17–19]. Collectively, these patterns indicate that what initially appears as a time-varying effect of the policy is more likely an artifact of the evolution of the at-risk patients which at the start were predominantly comprised of Status 1 and 2 patients.

We also observed a benefit among post-policy, low-acuity patients. Low-acuity candidates experienced increased or equivalent transplantation rates across landmark timepoints, and myocardial recovery and death/deterioration remained consistently more favorable. The relative proportions of Status 4-6 patients remained stable throughout the follow-up time and are likely to reflect a policy-era effect rather than compositional shifts of the low-acuity cohort. Together, these findings imply that the 2018 policy revision may have enhanced overall system efficiency and optimized organ access across the spectrum of patient acuity. As an example, the improved myocardial recovery post-policy may be due to greater opportunity for guideline directed medical therapy optimization during longer waitlist intervals under a policy allocation that favors longer waiting times for lower-acuity ambulatory patients [20, 21].

### Long-Distance Donor Utilization Increased Post-Policy Change and Declined over Time

Similar to prior literature, we observed that far-away donor utilization increased after policy implementation in both high- and low-acuity candidates, demonstrating that broader geographic sharing is an important feature of the current donor pool [4, 5]. Among high-acuity patients, the use of donor hearts procured ≥500 NM was highest shortly after listing and declined as early transplants were completed. This front-loaded pattern corresponds to the rapid removal of Status 1 and 2 candidates soon after listing. Low-acuity candidates also experienced increased access to far-away donors, but the magnitude of use was smaller early in the waitlist course and remained stable over time. These patterns suggest that broader geographic sharing facilitates early access for the sickest candidates while also broadening the donor pool for patients of lower acuity or who remain on the waitlist longer.

Our findings add temporal nuance to the literature evaluating the 2018 policy change [2, 4–13, 17–19]. They affirm that the policy’s intended goals of rapid heart allocation to the sickest patients and a reduction in waitlist mortality were achieved early after waitlist listing. They also highlight that broader geographic sharing may be an important factor in the sustained improvements for low-acuity patients. Lastly, the long-distance donor use patterns highlight potential opportunities for continued integration of organ preservation technologies to further expand the donor pool [22–28].

### Limitations

This study has several limitations. First, the retrospective nature and reliance on registry data expose this analysis to potential data inaccuracies, missing values, and reporting biases. Second, unmeasured confounders, such as detailed clinical characteristics, center-specific clinical management strategies, regional variations in organ availability, transplantation practices, and waitlist dynamics were not captured. This includes how practices may have been altered during the height of the COVID-19 pandemic. Third, we were unable to assess how donor pool growth, including the increasing use of donation-after-cardiac-death donors, may have influenced findings. Related, our use of the UNOS “delisted for myocardial recovery” code as a proxy for functional recovery has limitations in that no data is available following delisting. As such, post-delisting mortality and/or morbidity is unknown, possibly skewing the final recovery signal findings. Finally, harmonizing acuity across policy eras is imperfect as the mapping of old Status 1A to the current Status 1-3 does not fully account for concurrent practice developments such as increased temporary mechanical circulatory support (tMCS) utilization, axillary IABP use, Impella 5.5 availability, or changes in donor acceptance practices brought about following the policy change. Similarly, we could not account for heterogeneity in the acuity groups, such as HF etiology (hypertrophic cardiomyopathy, amyloidosis, tMCS, or LVAD) at the time of listing. We acknowledge that these status designations contain heterogeneous risk profiles, an issue the 2018 policy sought to address. Comparing outcomes in the first 1-2 years following the policy change to the remaining post-2018 period or period following the aforementioned clinical developments would provide valuable further insight into the evolving temporal landscape beyond the policy change itself. Nevertheless, comparing new Status 1-3 to old Status 1A (and similarly aligning lower-acuity statuses) remains the most conceptually consistent way to evaluate analogous levels of medical urgency before and after the policy change. As such, our approach may not fully capture within-status variability, and differences observed between eras should be interpreted accordingly.

## 5.0 Conclusion

The 2018 UNOS heart allocation policy produced meaningful improvements in waitlist outcomes, with a concentrated early transplantation benefit among high-acuity patients and advantages across all waitlist outcomes for low-acuity candidates. Temporal patterns in transplantation hazards reflect rapid early removal of the sickest patients and early peak use of far-away donors rather than intrinsic time-dependent policy effects. These findings highlight the effectiveness of broader geographic sharing while underscoring the lack of true time-based effects of the new waitlist system as the field advances toward continuous allocation and future organ-distribution and preservation innovations. The anticipated continuous allocation framework should consider how patients move through the waitlist in light of broader access geographic sharing and novel organ preservation technology to meet the goals of minimizing waitlist mortality and maximizing organ allocation to the sickest patients.

## Data Availability

All data produced in the present study are available upon reasonable request to the authors.

## Acknowledgements

All authors have read and approved the submitted manuscript and mention of their names. All listed authors meet the ICMJE criteria for authorship via the following:

1. *Study conception and design: AMW, MSC, JF, ASV*
2. *Analysis and interpretation of data: AMW, MSC, JK, MWF, JN, MS, ESK*
3. *Drafting of the manuscript submitted: AMW, MSC, JF, JK, MWF, JN, MS, ESK, ASV*
4. *Agreement to be accountable for all aspects of the work ensuring that questions related to the accuracy or integrity of any part of the work are appropriately investigated and resolved: AMW, MSC, JF, JK, MWF, JN, MS, ESK, ASV*

## Non-standard Abbreviations and Acronyms

BMI: body mass index
NM: nautical miles
IQR: interquartile range
sdHR: sub-distribution hazard ratio
csHR: cause-specific hazard ratio
UNOS: United Network for Organ Sharing
LVAD: left ventricular assist device
VA-ECMO: veno-arterial extracorporeal membrane oxygenation
IABP: intra-aortic balloon pump
ICD: implantable cardioverter-defibrillator
tMCS: temporary mechanical circulatory support
HF: heart failure

