## Supplemental Tables 1, 2, 3 for "United Network for Organ Sharing Policy Change Impact on Competing Risks in Heart Transplantation: Landmark Analysis of Waitlist Outcomes over Time"

**Table of Contents:**

1. Supplemental Table 1: Full results of the competing risks landmark analysis for each waitlist outcome of interest in high-acuity patients—Page 2, Line 9
2. Supplemental Table 2: Full results of the competing risks landmark analysis for each waitlist outcome of interest in low-acuity patients—Page 3, Line 12
3. Supplemental Table 3: Total count and overall proportion of "far-away" donor (≥500 NM from recipient) usage stratified by policy era and listing status—Page 4, Line 15

Supplemental Table 1

**High-acuity**

| | sdHR | | | | | | csHR | | | | | |
|---|---|---|---|---|---|---|---|---|---|---|---|---|
| | Heart Transplant | | Death/Deterioration | | Myocardial Recovery | | Heart Transplant | | Death/Deterioration | | Myocardial Recovery | |
| **Weeks** | HR 95% CI | p-value | HR 95% CI | p-value | HR 95% CI | p-value | HR 95% CI | p-value | HR 95% CI | p-value | HR 95% CI | p-value |
| **0** | 2.13 (2.04, 2.22) | <0.001 | 0.41 (0.37, 0.46) | <0.001 | 0.42 (0.29, 0.61) | <0.001 | 2.29 (2.19, 2.38) | <0.001 | 0.96 (0.86, 1.07) | 0.454 | 1.38 (0.94, 2.02) | 0.097 |
| **1** | 1.81 (1.73, 1.89) | <0.001 | 0.52 (0.47, 0.59) | <0.001 | 0.70 (0.46, 1.07) | 0.10 | 1.92 (1.84, 2.01) | <0.001 | 0.98 (0.87, 1.10) | 0.70 | 1.56 (1.02, 2.39) | 0.04 |
| **2** | 1.57 (1.5, 1.65) | <0.001 | 0.62 (0.55, 0.71) | <0.001 | 0.89 (0.56, 1.41) | 0.62 | 1.67 (1.59, 1.75) | <0.001 | 1.03 (0.91, 1.17) | 0.632 | 1.66 (1.05, 2.63) | 0.030 |
| **4** | 1.38 (1.3, 1.46) | <0.001 | 0.68 (0.59, 0.79) | <0.001 | 1.17 (0.71, 1.91) | 0.53 | 1.42 (1.34, 1.51) | <0.001 | 0.97 (0.84, 1.11) | 0.628 | 1.68 (1.03, 2.73) | 0.038 |
| **8** | 1.18 (1.09, 1.27) | <0.001 | 0.76 (0.63, 0.91) | 0.0025 | 1.24 (0.72, 2.14) | 0.43 | 1.17 (1.08, 1.26) | <0.001 | 0.9 (0.75, 1.08) | 0.271 | 1.34 (0.77, 2.31) | 0.296 |
| **16** | 0.89 (0.81, 0.99) | 0.027 | 0.86 (0.68, 1.07) | 0.18 | 1.11 (0.59, 2.1) | 0.74 | 0.87 (0.79, 0.96) | 0.0052 | 0.78 (0.62, 0.98) | 0.030 | 0.88 (0.46, 1.69) | 0.693 |
| **24** | 0.81 (0.72, 0.91) | <0.001 | 1.01 (0.78, 1.31) | 0.93 | 0.99 (0.48, 2.02) | 0.98 | 0.79 (0.70, 0.89) | <0.001 | 0.86 (0.67, 1.12) | 0.271 | 0.75 (0.36, 1.55) | 0.438 |

Supplemental Table 2

**Low-acuity**

| | sdHR | | | | | | csHR | | | | | |
|---|---|---|---|---|---|---|---|---|---|---|---|---|
| | Heart Transplant | | Death/Deterioration | | Myocardial Recovery | | Heart Transplant | | Death/Deterioration | | Myocardial Recovery | |
| **Weeks** | HR 95% CI | p-value | HR 95% CI | p-value | HR 95% CI | p-value | HR 95% CI | p-value | HR 95% CI | p-value | HR 95% CI | p-value |
| **0** | 1.30 (1.26, 1.35) | <0.001 | 0.56 (0.51, 0.61) | <0.001 | 1.32 (1.17, 1.5) | <0.001 | 1.27 (1.23, 1.31) | <0.001 | 0.66 (0.6, 0.71) | <0.001 | 1.57 (1.38, 1.79) | <0.001 |
| **1** | 1.25 (1.21, 1.30) | <0.001 | 0.58 (0.54, 0.64) | <0.001 | 1.35 (1.18, 1.53) | <0.001 | 1.22 (1.18, 1.26) | <0.001 | 0.66 (0.61, 0.72) | <0.001 | 1.53 (1.35, 1.74) | <0.001 |
| **2** | 1.22 (1.18, 1.27) | <0.001 | 0.59 (0.54, 0.64) | <0.001 | 1.38 (1.22, 1.57) | <0.001 | 1.19 (1.14, 1.23) | <0.001 | 0.66 (0.6, 0.72) | <0.001 | 1.53 (1.35, 1.73) | <0.001 |
| **4** | 1.16 (1.11, 1.2) | <0.001 | 0.62 (0.56, 0.68) | <0.001 | 1.43 (1.26, 1.62) | <0.001 | 1.12 (1.08, 1.17) | <0.001 | 0.67 (0.61, 0.73) | <0.001 | 1.52 (1.34, 1.72) | <0.001 |
| **8** | 1.07 (1.03, 1.12) | 0.001 | 0.65 (0.6, 0.72) | <0.001 | 1.53 (1.35, 1.74) | <0.001 | 1.05 (1.01, 1.09) | 0.027 | 0.68 (0.62, 0.75) | <0.001 | 1.55 (1.36, 1.75) | <0.001 |
| **16** | 0.97 (0.92, 1.01) | 0.180 | 0.72 (0.65, 0.8) | <0.001 | 1.65 (1.45, 1.87) | <0.001 | 0.96 (0.92, 1.01) | 0.112 | 0.72 (0.65, 0.8) | <0.001 | 1.59 (1.4, 1.81) | <0.001 |
| **24** | 0.94 (0.89, 0.99) | 0.022 | 0.69 (0.62, 0.77) | <0.001 | 1.68 (1.47, 1.93) | <0.001 | 0.93 (0.88, 0.98) | 0.008 | 0.67 (0.6, 0.75) | <0.001 | 1.6 (1.4, 1.83) | <0.001 |

Supplemental Table 3

| Era | Trend Test (p-value) | Landmark Week | Listing Status | Count (≥500 NM) | Total Transplants | Percent (≥500 NM) |
|---|---|---|---|---|---|---|
| Pre-policy | <0.001 | 1 | Status 1A | 47 | 576 | 8.20% |
| | | 2 | | 31 | 479 | 6.50% |
| | | 4 | | 42 | 820 | 5.10% |
| | | 8 | | 36 | 1026 | 3.50% |
| | | 16 | | 37 | 1044 | 3.50% |
| | | 24 | | 27 | 735 | 3.70% |
| Post-policy | 0.0145 | 1 | Status 1 | 58 | 728 | 8.00% |
| | | 2 | | 20 | 240 | 8.30% |
| | | 4 | | 7 | 183 | 3.80% |
| | | 8 | | 7 | 162 | 4.30% |
| | | 16 | | 1 | 86 | 1.20% |
| | | 24 | | 3 | 29 | 10.30% |
| | <0.001 | 1 | Status 2 | 350 | 1878 | 18.60% |
| | | 2 | | 189 | 1283 | 14.70% |
| | | 4 | | 135 | 1283 | 10.50% |
| | | 8 | | 98 | 1015 | 9.70% |
| | | 16 | | 75 | 737 | 10.20% |

| | | | | | | |
|---|---|---|---|---|---|---|
| | | 24 | | 29 | 299 | 9.70% |
| | 0.005 | 1 | Status 3 | 83 | 351 | 23.60% |
| | | 2 | | 53 | 253 | 20.90% |
| | | 4 | | 69 | 316 | 21.80% |
| | | 8 | | 60 | 306 | 19.60% |
| | | 16 | | 47 | 301 | 15.60% |
| | | 24 | | 26 | 162 | 16.00% |
